# Structural and neurophysiological alterations in Parkinson’s disease are aligned with cortical neurochemical systems

**DOI:** 10.1101/2023.04.04.23288137

**Authors:** Alex I. Wiesman, Jason da Silva Castanheira, Edward A. Fon, Sylvain Baillet, PREVENT-AD Research Group, Quebec Parkinson Network

**Author notes:** Corresponding author <u>Correspondence</u>: Alex I. Wiesman, PhD, McConnell Brain Imaging Centre, Montreal Neurological Institute, McGill University, Montreal QC. Data used in preparation of this article were obtained from the Pre-symptomatic Evaluation of Novel or Experimental Treatments for Alzheimer’s Disease (PREVENT-AD) program (https://douglas.research.mcgill.ca/stop-ad-centre), data release 6.0. A complete listing of PREVENT-AD Research Group can be found in the PREVENT-AD database: https://preventad.loris.ca/acknowledgements/acknowledgements.php?date=[2022-02-01]. The investigators of the PREVENT-AD program contributed to the design and implementation of PREVENT-AD and/or provided data but did not participate in analysis or writing of this report.

## Abstract

Parkinson’s disease (PD) affects cortical structures and neurophysiology. How these deviations from normative variants relate to the neurochemical systems of the cortex in a manner corresponding to motor and cognitive symptoms is unknown. We measured cortical thickness and spectral neurophysiological alterations from structural magnetic resonance imaging and task-free magnetoencephalography in patients with idiopathic PD (N_MEG_ = 79; N_MRI_ = 65), contrasted with similar data from matched healthy controls (N_MEG_ = 65; N_MRI_ = 37). Using linear mixed-effects models and cortical atlases of 19 neurochemical systems, we found that the structural and neurophysiological alterations of PD align with several receptor and transporter systems (acetylcholine, serotonin, glutamate, and noradrenaline) albeit with different implications for motor and non-motor symptoms. Some neurophysiological alignments are protective of cognitive functions: the alignment of broadband power increases with acetylcholinergic systems is related to better attention function. However, neurochemical alignment with structural and other neurophysiological alterations is associated with motor and psychiatric impairments, respectively. Collectively, the present data advance understanding of the association between the nature of neurophysiological and structural cortical alterations in PD and the symptoms that are characteristic of the disease. They also demonstrate the value of a new nested atlas modeling approach to advance research on neurological and neuropsychiatric diseases.

## Introduction

Parkinson’s disease (PD) is a progressive neurodegenerative disorder that impairs movement and cognition^1^, with frequent neuropsychiatric complications^2^. Movement features of PD include bradykinesia, muscular rigidity, and resting tremor, while cognitive declines are most common in the domains of attention and working memory. Alongside characteristic degeneration of neurons in deep brain nuclei, patients with PD present with functional and structural alterations at the level of the cortex. Broad changes in neurophysiological brain activity across the frequency spectrum^3,4^ are accompanied by the structural thinning of frontal and posterior cortices^5-7^. These effects scale in magnitude with symptoms^4,6,8-13^, indicating their relevance to clinical monitoring and disease-modifying intervention.

Which cortical regions are primarily affected by structural and neurophysiological changes in PD, and how they align with the cortical gradients of the functional hierarchy is clinically meaningful^10,13,14^. How these alterations might relate to the relative concentration of major neurochemical systems of the cortex is key to understanding their mechanistic underpinnings and the definition of novel therapeutic targets.

Motor dysfunction in PD arises from a complex combination of neurochemical changes^17^, including cell death of dopaminergic neurons in deep brain nuclei^14^, as well as cortical alterations of the serotongergic^15,16^ and noradrenergic^17-20^ systems. Cognition is also affected along the PD continuum, with attentional deficits being of core clinical significance because they increase the risk of falls^21,22^ and negatively affect quality of life^23^. Attention is a multifaceted cognitive area subserved by multiple cortical neurotransmitter systems, with essential contributions from the acetylcholine and norepinephrine systems^24-26^. These latter are both aberrant in PD^18,22,27^, and represent emerging targets for pharmacotherapy^18,28^.

Here, we systematically examine in patients with idiopathic PD the alignment of disease-related cortical alterations with the principal neurochemical gradients of the cortex, and investigate its protective and adverse effects on symptoms.^29^

## Results

### Principal Spatial Gradients of Normative Neurotransmitter System Density

We first reduced the 19 normative atlases of neurotransmitter system densities (Figure S1) into their principal components of spatial variation across the cortex, four of which were significant (1,000 permutations; 81.7% of total spatial variance; Figure 1). The first gradient (pc1; eigenvalue = 6.13; *p* < .001) accounted for 32.3% of the total spatial variation in normative neurotransmitter systems and highlighted cortical regions with higher density of most modeled receptors and transporters, with the exception of GABAergic and several serotonergic systems. As reported previously,^29^ this gradient shared significant spatial variability with synaptic density (i.e., glycoprotein; *r* = .41, *p*_*spin*_ < .001). The second gradient (pc2; eigenvalue = 4.79; *p* < .001) accounted for 25.2% of total variation, and highlighted the divergence of cortical regions rich in norepinephrine, GABA, acetylcholine, and 5-HT1b/5-HT6 serotonin systems from those with higher densities of dopamine, mu-opioid, cannabinoid, and other serotonergic (5-HT1a, 5-HT4, and 5-HTT) systems. The third gradient (pc3; eigenvalue = 2.56; *p* < .001) explained 13.5% of the total variance, and described a spatial pattern of relative increase in GABAergic, serotonergic (5-HT2a and 5-HTT), and glutamatergic (NMDA) systems versus decreased mu-opioid and cannabinoid systems. Finally, the fourth gradient (pc4; eigenvalue = 2.04; *p* < .001) accounted for 10.7% of the total variance and mapped onto regions with low serotonergic (5-HT2a and 5-HT4) and acetylcholinergic (M1) receptor densities.

**Figure 1.**
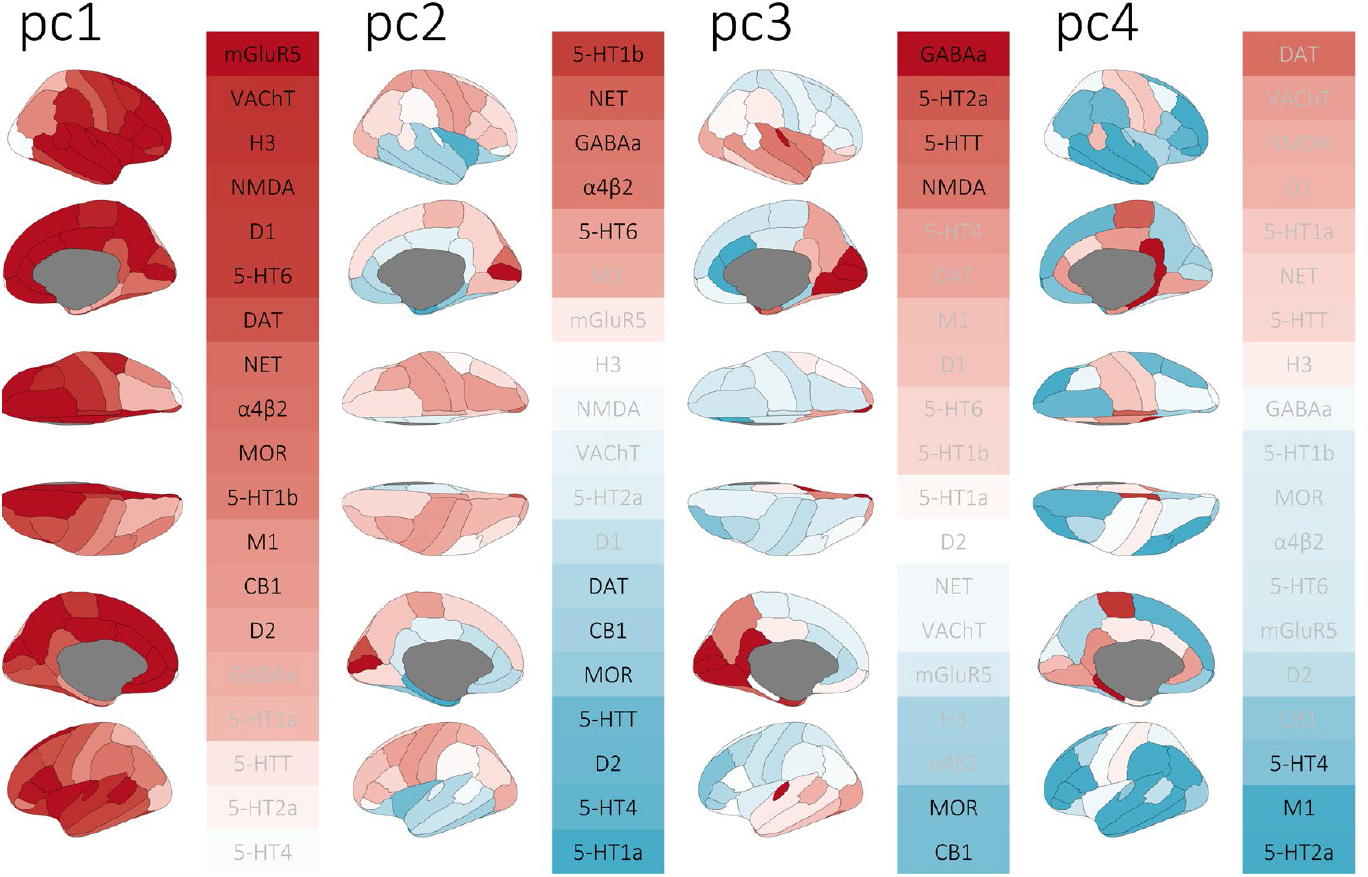
Normative gradients of cortical neurotransmitter system density derived from 19 normative neurotransmitter atlases using principal component analysis. The first four gradients, labeled from pc1 to pc4, explained >80% of the variance across atlases and are shown as cortical maps of their respective spatial principal components (all *p*’s < .001; 1,000 permutations). Heatmaps to the right of each cortical map show their respective loadings (i.e., eigenvectors) across the 19 normative atlases. Positive and negative loadings are colored red and blue, respectively, and labels in black indicate statistical significance *(p*’s < .05; 1,000 permutations).

### Alignment of Neurophysiological Alterations with Neurochemical Systems

The cortical maps of group means and variability of neurophysiological alterations derived from the present cohorts highlight decreased delta and beta activity in fronto-parietal and temporo-parietal cortices, and increased theta and alpha activity in temporo-parietal and frontal somato-motor regions, respectively, in patients with PD (Figure S2).

We tested whether these neurophysiological alterations align with the four normative neurochemical gradients (see Methods; Figure 2). We used nested linear models towards this goal, because they account for both the cross-cortical alignment of PD-related alterations with the neurochemical gradients and the inter-individual variability of these alignments across patients. This approach also then allows for control of nuisance covariates.

**Figure 2.**
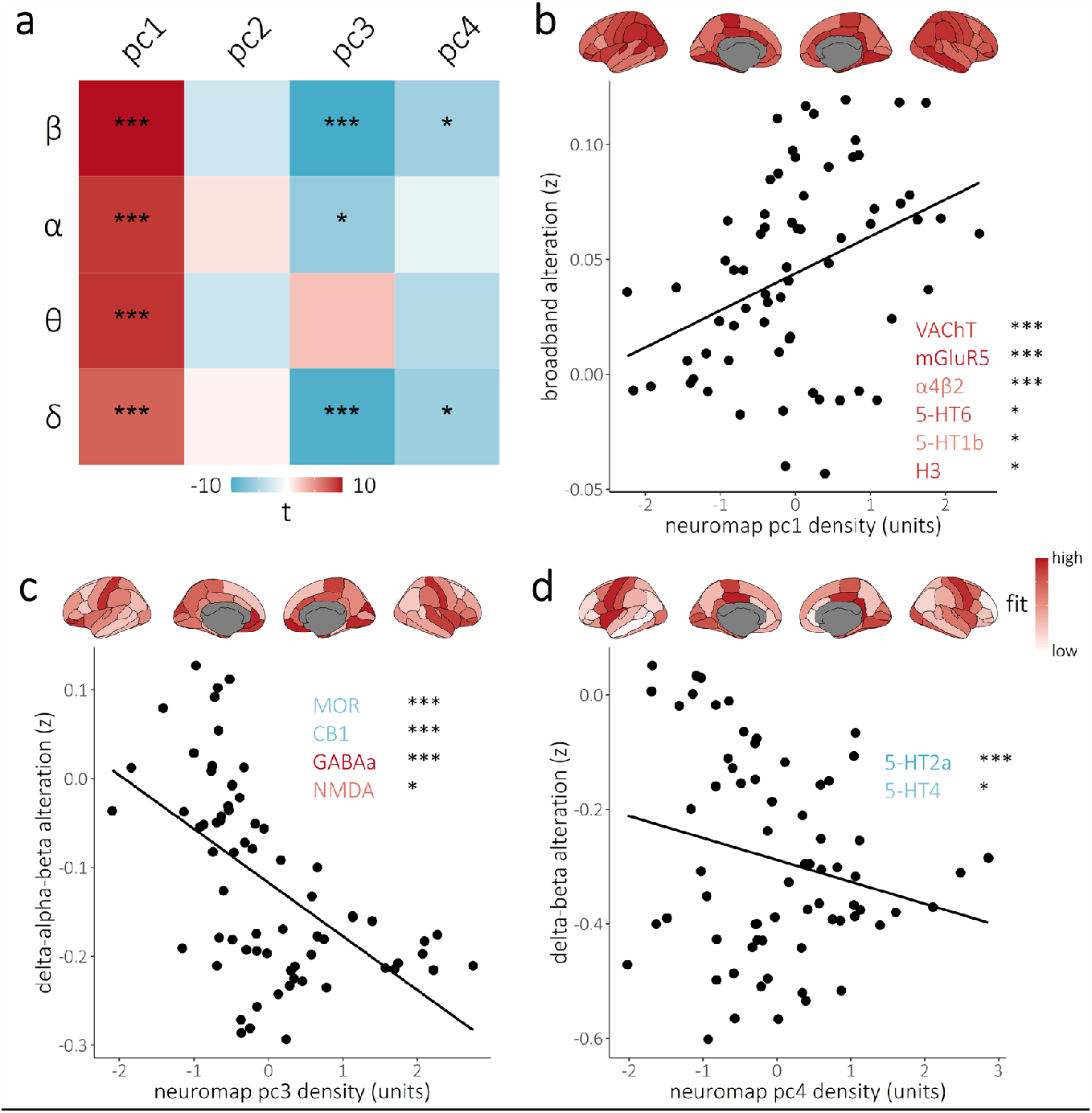
Neurophysiological alterations in Parkinson’s disease align with neurochemical systems. The heatmap in (a) indicates the strength of the alignment between frequency-specific neurophysiological alterations (y-axis) and the principal cortical gradients of normative neurotransmitter systems from Figure 1 (x-axis). Colors indicate the t-values of these relationships, with asterisks indicating statistical significance accounting for multiple comparisons. Scatterplots in (b-d) highlight the significant alignments from (a). Neurophysiological alterations exhibiting similar alignments with the same neurochemical gradient were averaged over frequency bands, and these values were then averaged across individuals per cortical region (y-axes) and plotted against neurotransmitter system principal gradients (x-axes) for visualization. The cortical maps above each scatter plot show the standardized contribution to the model fit of each cortical region across individuals (see *Methods: Modeling of Spatial Colocalization* for details), with darker colors indicating cortical regions where the alignment between neurophysiological alterations and neurochemical gradients is stronger. Inlaid neurotransmitter receptor/transporter abbreviations indicate the respective atlases that significantly align with the relevant alterations (y-axes), as per post-hoc testing, with the color and saturation of each label indicating the strength and sign, respectively, of the corresponding loading onto the neurochemical principal component. \*\*\**p*_*FDR*_ < .005, \**p*_*FDR*_ < .05.

We found that PD-related increases in broadband neurophysiological activity map to the first principal neurochemical gradient (pc1; delta: *t* = 3.45, *p*_*FDR*_ = .002; theta: *t* = 4.28, *p*_*FDR*_ < .001; alpha: *t* = 4.22, *p*_*FDR*_ < .001; beta: *t* = 4.69, *p*_*FDR*_ < .001), and specifically to the cortical distribution of acetylcholinergic (post-hoc tests; VAChT: *t* = 4.49, *p*_*FDR*_ < .001; α4β2: *t* = 3.14; *p*_*FDR*_ = .008), glutamatergic (mGluR5: *t* = 3.86; *p*_*FDR*_ < .001), serotonergic (5-HT6: *t* = 2.60; *p*_*FDR*_ = .028; 5-HT1b: *t* = 2.58; *p*_*FDR*_ = .028), and histamine (H3: *t* = 2.47; *p*_*FDR*_ = .032) systems. This association remained significant after controlling for the overlap of the first neurochemical gradient with regional synaptic density (*t* = 3.95, *p* < .001).

The cortical expression of neurophysiological alterations in the delta, alpha, and beta bands is also spatially aligned with the third neurochemical gradient (pc3; delta: *t* = -3.95, *p*_*FDR*_ < .001; alpha: *t* = -2.46, *p*_*FDR*_ = .031; beta: *t* = -4.12, *p*_*FDR*_ < .001): depressed delta, alpha, and beta activity in PD is more pronounced in regions with higher concentrations of GABAa (*t* = -3.15; *p*_*FDR*_ = .003) and NMDA (*t* = -2.52; *p*_*FDR*_ = .018) and lower concentrations of MOR (*t* = 4.65; *p*_*FDR*_ < .001) and CB1 (*t* = 4.27; *p*_*FDR*_ < .001) receptors. We also found associations between delta and beta activity and the fourth neurochemical gradient (pc4; delta: *t* = -2.26, *p*_*FDR*_ = .047; beta: *t* = -2.21, *p*_*FDR*_ = .048), such that depressed delta and beta neurophysiological activity in PD is more pronounced in cortical regions with lower levels of serotonergic receptors (5-HT2a: *t* = 3.33; *p*_*FDR*_ = .003; 5-HT4: *t* = 2.16; *p*_*FDR*_ = .047). All the reported spatial alignments remained significant (all *p*’s < .05) after adding additional nuisance covariates (beyond age) to the model, including head motion, eye movements, and heart rate variability.

We did not find significant spatial alignment between the neurochemical gradients and arrhythmic neurophysiological alterations in PD (pc1: *t* = 0.13, *p*_*FDR*_ = .901; pc2: *t* = 1.94, *p*_*FDR*_ = .226; pc3: *t* = -0.72, *p*_*FDR*_ = .846; pc4: *t* = 0.48, *p*_*FDR*_ = .846).

### Associations with Non-motor Symptoms of Parkinson’s Disease

We examined whether the alignment of neurophysiological alterations in PD and neurochemical gradients is moderated by domain-specific cognitive abilities, motor function, and self-reported psychiatric symptoms. We found that patients with stronger broadband neurophysiological alterations in cortical regions rich in acetylcholine, glutamate, serotonin, and histamine systems have preserved attention abilities (alignment with first neurochemical gradient; *t* = 3.81, *p*_*FDR*_ < .001; Figure 3a).

**Figure 3.**
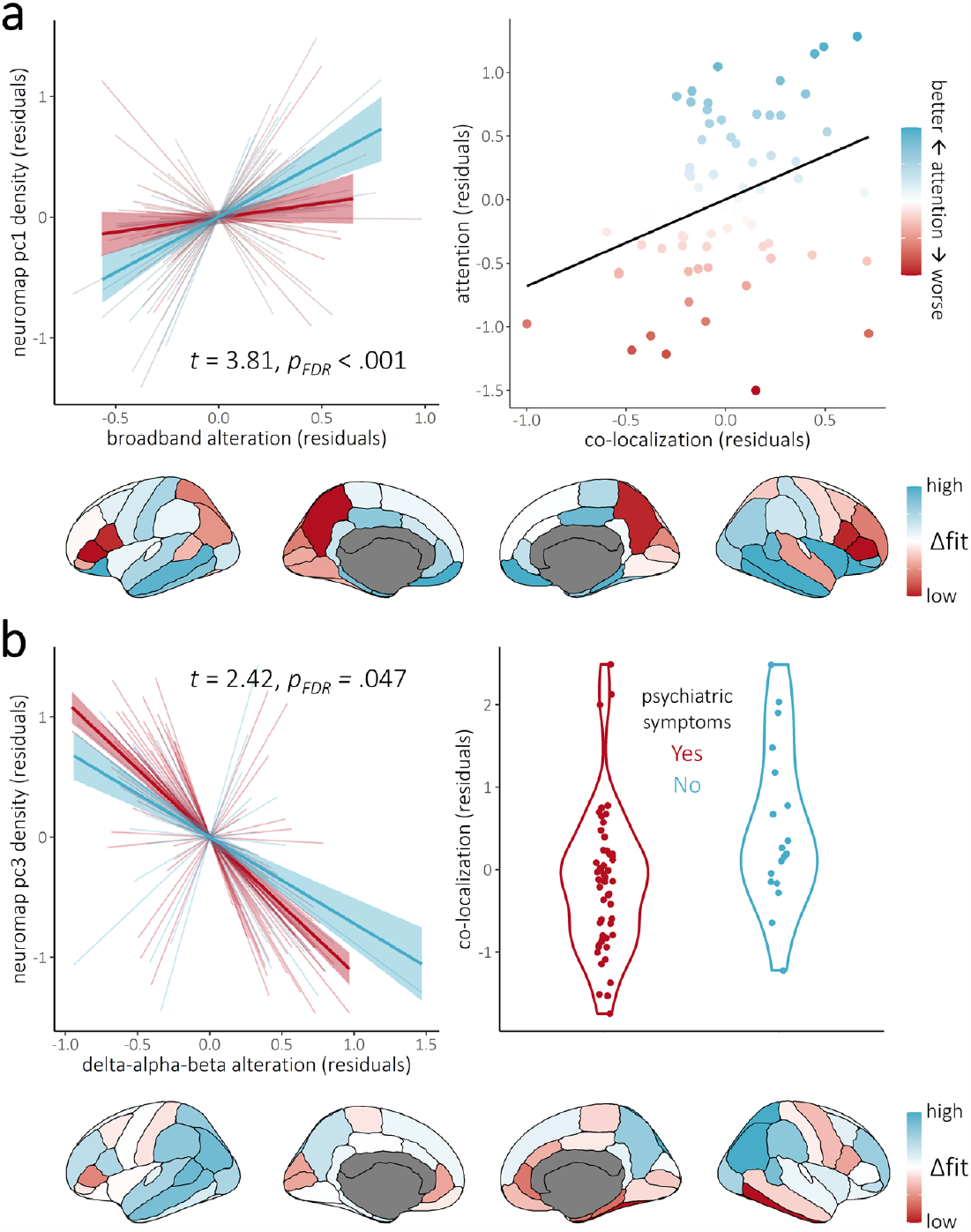
The alignment between Parkinson’s disease neurophysiological alterations and cortical neurochemical system densities relates to preserved attention abilities in patients. (a) We observe a moderation of the alignment between the first principal neurochemical gradient and Parkinson’s disease neurophysiological alterations by attention abilities: patients with stronger alignment had preserved attention abilities. Left: each line represents the alignment between neurophysiological alterations and the neurochemical gradient per patient with Parkinson’s disease (thinner lines with no shading), colored according to each patient’s composite scores on neuropsychological tests of attention. Overlaid thicker lines show this relationship across patients with attention scores in the top (blue) and bottom (red) quartiles. Shading indicates 95-% confidence intervals. Right: individual patient slopes are plotted (x-axis) against attention scores (y-axis and colors) to emphasize the continuum of the moderation effect. Bottom: brain maps of the regional difference in standardized model fit between patients with intact (top quartile) and impaired (bottom quartile) attention abilities. (b) We observe a similar moderation effect by psychiatric symptoms, such that the alignment of the third neurochemical gradient with delta-alpha-beta-band neurophysiological alterations is stronger in patients with self-reported manifestations of depression and/or anxiety.

The alignment between neurophysiological alterations in the delta, alpha, and beta bands and the third neurochemical gradient is moderated by the presence of self-reported psychiatric symptoms (e.g., depression and /or anxiety; *t* = 2.42, *p*_*FDR*_ = .047; Figure 3b): patients who experience psychiatric symptoms show more depressed neurophysiological activity in cortical regions rich in GABA and glutamate receptors. Conversely, patients who do not report psychiatric symptoms show stronger levels of neurophysiological alteration in cortical regions rich with mu-opioid and cannabinoid receptors.

No significant moderations were found by motor function of the alignment between neurophysiological alterations and neurochemical gradients (all *p*’s > .05).

These observations remained significant (all *p*’s < .05) after adding nuisance covariates (beyond age) to the model, including head motion, eye movements, and heart rate variability.

### Alignment of Structural Alterations with Neurochemical Systems and Association with Motor Symptoms of Parkinson’s Disease

The cortical maps of structural alterations in this patient cohort highlight thinning of temporo-parietal cortices (Figure S2). Our data show that these PD-related structural alterations align with the second neurochemical gradient (pc2; *t* = 2.93, *p*_*FDR*_ = .013; Figure 4a-b), such that cortical thinning is less pronounced in regions with high densities of norepinephrine (NET: *t* = 3.36; *p*_*FDR*_ = .010), serotonin (5-HT1b: *t* = 3.15; *p*_*FDR*_ = .010; 5-HT6: *t* = 2.63; *p*_*FDR*_ = .026), and acetylcholine (α4β2: *t* = 2.79; *p*_*FDR*_ = .021) transporters/receptors. The relative sparing of these regions from cortical thinning was associated with preserved motor function in patients (i.e., UPDRS-III scores; *t* = -2.15, *p* = .031; Figure 4c).

**Figure 4.**
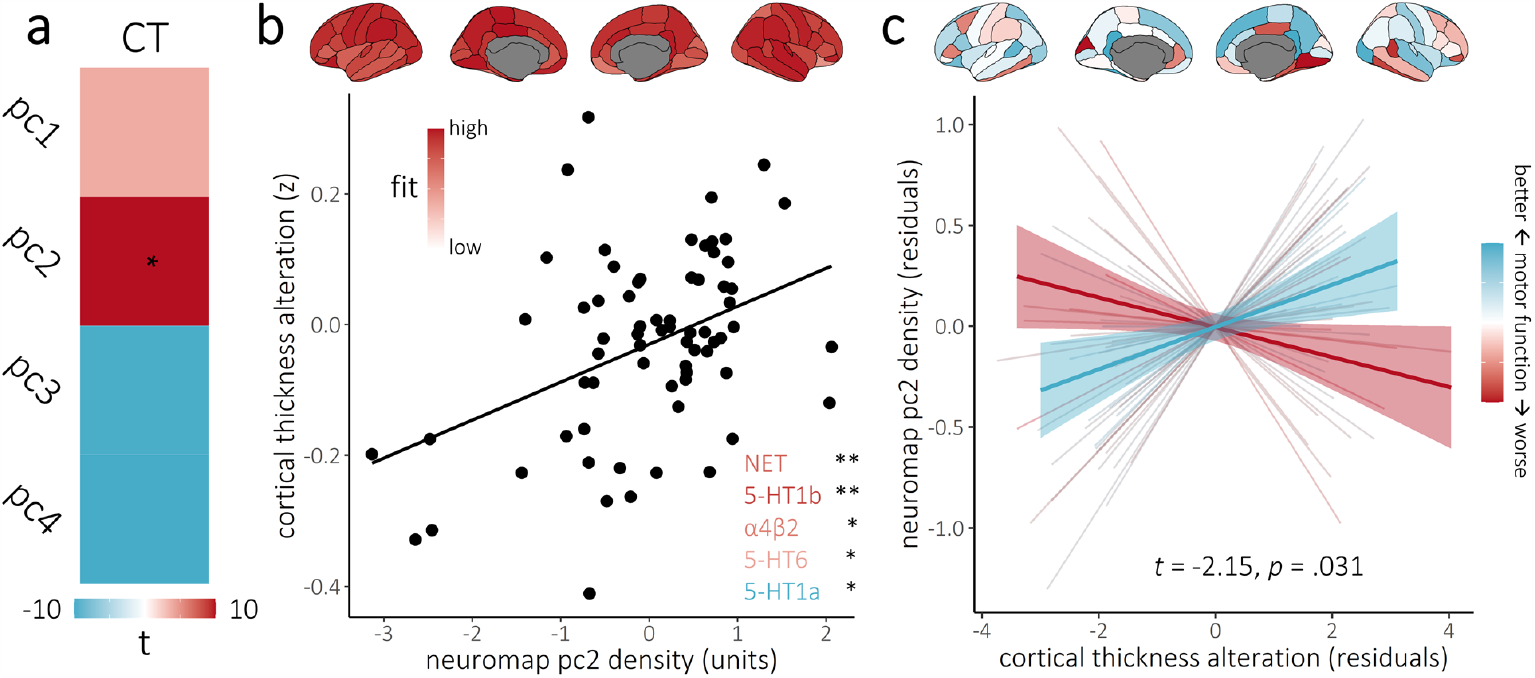
Cortical thickness alterations in Parkinson’s disease are aligned with neurochemical systems. The heatmap in (a) indicates the strength of the overlap between cortical thickness alterations (x-axis) and the principal cortical gradients of normative neurotransmitter systems from Figure 1 (y-axis). Colors indicate the t-values of these overlaps, with asterisks indicating statistical significance accounting for multiple comparisons. The scatterplot in (b) highlights the significant association from (a). Brain maps above represent the standardized contribution to the model fit of the respective cortical region across individuals. Inlaid neurotransmitter receptor/transporter abbreviations indicate the atlases significantly related to the relevant alterations (y-axis) in post-hoc testing, with the color and saturation of each label indicating the strength and sign, respectively, of its loading onto the principal component. (c) We observe a moderation of the alignment between the second principal neurochemical gradient and Parkinson’s disease cortical thickness alterations by motor function: patients with stronger alignment had preserved motor function. Each line represents the alignment between cortical thickness alterations and the neurochemical gradient per patient with Parkinson’s disease (thinner lines with no shading), colored according to each patient’s composite scores on clinical tests of motor function. Overlaid thicker lines show this relationship across patients with motor function scores in the top (blue) and bottom (red) quartiles. Shading indicates 95-% confidence intervals. Brain maps above show the regional difference in standardized model fit between patients with intact (top quartile) and impaired (bottom quartile) motor function. \*\**p*_*FDR*_ < .01, \*\**p*_*FDR*_ < .05.

These observations remained significant (*p* < .05) with additional nuisance covariates (beyond age), including head motion, eye movements, and heart rate variability.

## Discussion

Alterations in cortical structure and neurophysiological activity are well-documented in patients with Parkinson’s disease. How their magnitude relate to clinical impairments is also well-studied, but whether their cortical distributions are related to the neurochemical organization of the cortex, and whether these topographies are clinically meaningful have been unanswered questions in the field.

Here, we demonstrate that structural and neurophysiological alterations in the brains of patients with PD align with the principal neurochemical organization of the human cortex. We also show that, beyond the magnitude of PD alterations, their topographical alignment with neurochemical boundaries is clinically meaningful. Overall, these findings contextualize decades of neuroimaging research in PD, and point at potential new research directions for disease monitoring and pharmacotherapy approaches that consider the spatial distributions of cortical alterations, in addition to their magnitude.

We discovered three patterns of PD neurophysiological alterations that follow distinct neurochemical gradients. Stronger broadband neurophysiological alterations are expressed in frontal cortical regions rich in cholinergic, metabotropic glutamatergic, serotonergic, and histamine systems. The cholinergic systems (VAChT and α4β2) play a key role in PD symptomatology^30^: their denervation occurs early in the disease course^31^, and their progressive disintegration is thought to be the primary driver of non-motor deficits in PD^22,30^. For example, frontal and temporo-parietal changes in cholinergic systems are related to aberrant dopaminergic signaling in the striatum, but only cholinergic function differentiates between patients with and without dementia^32^. A previous MEG study showed increased neurophysiological activity after administration of a cholinesterase inhibitor in a small group of patients with PD^33^. Our findings indicate that, even without explicit pharmacological manipulation of cholinergic systems, increased frontal neurophysiological activity in PD is indeed related to cholinergic function.

The patients with greater neurophysiological-neurochemical alignment with cholinergic systems have preserved attention function, indicating a normalizing/compensatory effect^25^. The impact of cholinergic dysfunction on attentional changes in patients with PD^27,30^ contributes to risk of falls^22^ and decreased quality of life^23^. As such, neurophysiological mapping with MEG, and potentially EEG, might be a more practical and less-invasive approach than PET for measuring the development of attentional deficits along the disease trajectory. Because MEG directly measures post-synaptic currents and is immune to vascular confounds, it also represents a unique asset to conduct neuropharmacological studies, and especially cholinergic therapy studies in PD.

Decreases in neurophysiological activity in the delta, alpha, and beta bands are stronger in cortical regions rich in glutamatergic and GABAergic receptors, and weaker in mu-opioid and cannabinoid-rich areas. This alignment was stronger in patients who experienced psychiatric symptoms. Substantial evidence^34-36^ supports the role of mu-opioid and endocannabinoid systems in depression and anxiety, indicating that a shift towards decreases in activity of these systems (relative to GABAergic and glutamatergic systems) might represent a compensatory or neuroprotective effect against psychiatric symptoms in PD.

The fourth neurochemical gradient is also aligned with delta-beta neurophysiological alterations, but their concordance was not significantly related to the clinical factors tested in the present study. Other clinical symptoms of PD include psychosis^37^ and hallucinations^38^; more research is required to test their associations with neurophysiological-neurochemical alignments.

We did not find any neurophysiological-neurochemical alignment that scaled with motor function. However, the patient participants in the present study were all under a stable regimen of dopaminergic medications, which normalize neurophysiological activity related to the motor characteristics of PD^39,40^. We can therefore hypothesize that neurophysiological-neurochemical alignments related to motor functions may be detectable in the “off” medication state of patients with PD.

We also find that PD-related structural thinning of the cortex aligns with neurochemical boundaries: regions rich in noradrenergic, serotonergic (notably excepting 5-HT1a), and cholinergic systems are generally spared from degeneration. Here, we did find that such selective sparing relates to preserved motor function. The neurotransmitter systems aligned with structural alterations are all impacted by PD^16,18,19,30,41,42^. Norepinephrine is produced by the locus coeruleus, which accumulates proteinopathy and is affected by degeneration in patients with PD^43^. Inhibition of norepinephrine reuptake improves motor learning in healthy adults^44^. Serotonin is also a potent modulator of several cortical neurochemical systems and brain networks^45,46^. However, noradrenergic, serotonergic, and cholinergic dysfunction are more often related to cognitive than motor deficits in PD^22,30,32,41,47-49^. One potential reason for this idiosyncrasy is that cognitive and somatomotor cortical systems interact extensively in the healthy brain^50-54^, and breakdown of these interactions leads to worsened performance on motor tests and higher risk of debilitating falls in PD^21,22,55,56^. This indicates that the moderation of structural-neurochemical alignments by motor abilities reported herein is multifaceted and may be related to processes higher in the hierarchy of motor functions. Future research with neuroimaging and motor tasks would help clarify these associations.

Collectively, our data show that the neurophysiological and structural brain changes in PD are aligned with specific neurochemical boundaries of the cortex, with distinctive relevance for cognitive/psychiatric and motor abilities. Specifically, we find that the neurochemical organization of neurophysiological alterations relates to cognitive impairments and psychiatric symptoms of the disease, and that motor impairments are related to the alignment between the topographies of structural degeneration and neurochemical systems.

From these observations, we can hypothesize that because neuromodulatory or pharmacological interventions against PD target neurophysiological processes, they are more likely to ameliorate cognitive impairments and reduce psychiatric symptoms, than they are to alleviate dopamine-resistant motor deficits. We anticipate a new vein of research questions may spawn from the present data, with respect to potential interactive effects of cortical cholinergic, serotonergic, and noradrenergic systems on structural and neurophysiological alterations in PD, and how the proposed atlas-based approach incorporating inter-individual variability can contribute to clinical neuroscience investigations. Importantly, this approach is applicable to any clinical disorder associated with cortical alterations detectable with neuroimaging.

## Materials & Methods

### Participants

The Research Ethics Board at the Montreal Neurological Institute reviewed and approved this study. Written informed consent was obtained from every participant following detailed description of the study, and all research protocols complied with the Declaration of Helsinki. Exclusionary criteria for all participants included current neurological (other than PD) or major psychiatric disorder; MEG contraindications; and unusable MEG or demographic data.

Healthy participants and patients with mild to moderate (Hoehn and Yahr scale: 1 – 3) idiopathic PD were enrolled in the Quebec Parkinson Network (QPN; https://rpq-qpn.ca/)^57^ initiative, which comprises extensive clinical, neuroimaging, neuropsychological, and biological profiling of participants. A sample of 79 patients with PD fulfilled the inclusion criteria. All patients with PD were prescribed a stable dosage of antiparkinsonian medication with satisfactory clinical response prior to study enrollment. Patients were instructed to take their medication as prescribed before research visits, and thus all data were collected in the practically-defined “ON” state.

Magnetoencephalography^58^ data from 65 healthy older adults were collated from the Quebec Parkinson Network (N = 10)^57^, PREVENT-AD (N = 40)^59^ and Open MEG Archive (OMEGA; N = 15)^60^ data repositories for comparative normalization of the MEG data from patients with PD. These participants were selected so that their demographic characteristics, including age (Mann-Whitney U test; *W* = 2349.50, *p* = .382), self-reported sex (chi-squared test; χ^2^ = 0.65, *p* = .422), handedness (chi-squared test; χ^2^ = 0.25, *p* = .883), and highest level of education (Mann-Whitney U test; *W* =2502.50, *p* = .444), did not statistically differ from those of the patient group. These participants underwent the same MEG data collection procedure using the same instrument as the patient group.

Structural T1 MRI data from 37 healthy older adults from the Quebec Parkinson Network were used for comparison with the statistics of T1 MRI data from patients with PD. These participants underwent the same T1 MRI data collection procedure using the same instrument as the 65 patients with Parkinson’s disease with MRI data. Once again, these participants were selected so that the group demographic characteristics, including age (Mann-Whitney U test; *W* = 1267.00, *p* = .656), self-reported sex (chi-squared test; χ^2^ = 3.59, *p* = .058), handedness (chi-squared test; χ^2^ = 0.29, *p* = .593), and highest level of education (Mann-Whitney U test; *W* = 1008.50, *p* = .395), did not significantly differ from those of the patient group.

Group demographic summary statistics and comparisons, as well as clinical summary statistics for the patient group, are provided in Tables S1-2.

### Clinical & Neuropsychological Testing

Standard clinical assessments were available for most of the patients with PD, including data regarding gross motor impairment (Unified Parkinson’s Disease Rating Scale – part III [UPDRS-III]; N = 61)^61^, general cognitive function (Montreal Cognitive Assessment [MoCA]; N = 70)^62^, and presence of self-reported psychiatric symptoms (binary response; N = 72). As described previously^10^, detailed neuropsychological data were also available for 69 patients with PD, and were used to derive composite scores across five domains: attention, executive function, visuospatial function, memory, and language.

### Magnetoencephalography Data Collection and Analyses

Eyes-open resting-state MEG data for 144 participants (79 patients with PD and 65 healthy controls) were collected using a 275-channel whole-head CTF system (Port Coquitlam, British Columbia, Canada) at a sampling rate of 2400 Hz and with an antialiasing filter with a 600 Hz cut-off. Noise-cancellation was applied using CTF’s software-based built-in third-order spatial gradient noise filters. Recordings lasted a minimum of 5 min^63^ and were conducted with participants in the upright position as they fixated a centrally-presented crosshair. The participants were monitored during data acquisition via real-time audio-video feeds from inside the MEG shielded room, and continuous head position was recorded during all sessions.

MEG preprocessing was performed with *Brainstorm*^64^ unless otherwise specified, with default parameters and following good-practice guidelines^65^. The data were bandpass filtered between 1–200 Hz to reduce slow-wave drift and high-frequency noise, and notch filters were applied at the line-in frequency and harmonics (i.e., 60, 120 & 180 Hz). Signal space projectors (SSPs) were derived around cardiac and eye-blink events detected from ECG and EOG channels using the automated procedure available in *Brainstorm*^66^, reviewed and manually-corrected where necessary, and applied to the data. Additional SSPs were also used to attenuate stereotyped artifacts on an individual basis. Artifact-reduced MEG data were then epoched into non-overlapping 6-second blocks and downsampled to 600 Hz. Data segments still containing major artifacts (e.g., SQUID jumps) were excluded for each session on the basis of the union of two standardized thresholds of ± 3 median absolute deviations from the median: one for signal amplitude and one for its numerical gradient. An average of 79.72 (SD = 13.82) epochs were used for further analysis (patients: 83.78 [SD = 7.24]; controls: 74.77 [SD = 17.82]) and the percent of epochs rejected did not differ between the groups (*p* = .123). Empty-room recordings lasting at least 2 minutes were collected on or near the same day as the participants’ visits and were processed using the same pipeline, with the exception of the artifact SSPs, to model environmental noise statistics for source mapping.

MEG data were coregistered to each individual’s segmented T1-weighted MRI (*Freesurfer recon-all*)^67^ using approximately 100 digitized head points. For participants with useable MEG but not MRI data (N = 14 patients with PD; N = 3 healthy older adults), we produced an individualized template with *Brainstorm*, by warping the default *Freesurfer* anatomy to the participant’s head digitization points and anatomical landmarks^68^. We produced source maps of the MEG sensor data with overlapping-spheres head models (15,000 cortical vertices, with current flows of unconstrained orientation) and the dynamic statistical parametric mapping (dSPM) approach, all with Brainstorm default parameter values, and informed by estimates of sensor noise covariance derived from the empty-room MEG recordings.

We obtained vertex-wise estimates of power spectrum density (PSD) from the source-imaged MEG data using Welch’s method (3-s time windows with 50% overlap), which we normalized to the total power of the frequency spectrum at each cortical location. These PSD data were next averaged over all artifact-free 6-second epochs for each participant, and over canonical frequency bands (delta: 2–4 Hz; theta: 5–7 Hz; alpha: 8–12 Hz; beta: 15–29 Hz).

We also extracted several metrics to test for potential confounds of our primary effects of interest. To determine whether topographical alignment effects between the neurophysiological alterations and the neurochemistry maps were related to broadband arrhythmic activity, we processed the PSDs with *specparam* (Brainstorm MATLAB version; frequency range = 2–40 Hz; Gaussian peak model; peak width limits = 0.5 –12 Hz; maximum n peaks = 3; minimum peak height = 3 dB; proximity threshold = 2 standard deviations of the largest peak; fixed aperiodic; no guess weight)^69^ to estimate the slope of the aperiodic component of the neurophysiological power spectrum. The resulting slope values were averaged within each region of the Desikan-Killiany atlas and normalized to comparable data from the healthy controls. To investigate the possible confounding effects of head motion, eye movements, and heart-rate variability, we extracted the root-sum-square of the reference signals from the MEG head position indicators, EOG, and ECG channels, respectively.

### Structural MRI Data Collection and Analyses

After visual inspection for excessive motion or other artifacts, cortical thickness estimates were derived from T1-weighted MRI data for 102 participants (65 patients with PD and 37 healthy controls) from a 3-Tesla Siemens Prisma scanner with a 32-channel head coil and the following sequences and parameters: 3D T1 Magnetization Prepared – Rapid Gradient Echo (MPRAGE); TR: 2300 ms; TE: 2.98 ms; flip angle: 9°; field of view: 256 mm; slice thickness: 1 mm. Cortical thickness was estimated using *FreeSurfer*^*67*^ (version 6.0.0; Linux centos6_x86_64) on a workstation running Ubuntu 18.04.6 LTS.

Structural T1 MRI data for an additional 55 healthy control participants (40 from PREVENT-AD and 15 from OMEGA) were used for MEG source mapping. These data were not included in the analysis of cortical thickness, as they were collected using different parameters and/or scanners than the data collected from patients with PD. Such differences do not meaningfully impact the estimation of MEG source images^70,71^, but are more likely to bias group-wise comparisons of cortical thickness.

### Normative Atlases of Neurotransmitter System Density

To circumvent the safety limitations of repeated positron emission tomography (PET) imaging, an open database of normative atlases of human neurotransmitter system densities has been developed: *neuromaps*^72^. This multi-atlas aggregates PET data for 19 different neurotransmitter receptors and transporters from non-overlapping participant samples comprising more than 1,200 different healthy individuals, and provides a key resource for interpreting the topography of cortical changes in clinical disorders through a neurochemical lens.

We used *neuromaps*^72^ to obtain mean cortical distribution maps of 19 receptors and transporters, following previously-established procedures^29^ and parcellated the resulting topographies according to the Desikan-Killiany atlas^73^. We obtained normative densities for dopamine (D1: 13 adults, [11C]SCH23390 PET; D2: 92, [11C]FLB-457, DAT: 174, [123I]-FP-CIT), serotonin (5-HT1a: 36, [11C]WAY-100635; 5-HT1b: 88, [11C]P943; 5-HT2a: 29, [11C]Cimbi-36; 5-HT4: 59, [11C]SB207145; 5-HT6: 30, [11C]GSK215083; 5-HTT: 100, [11C]DASB), acetylcholine (α4β2: 30, [18F]flubatine; M1: 24, [11C]LSN3172176; VAChT: 30, [18F]FEOBV), GABA (GABAa: 16, [11C]flumazenil), glutamate (NMDA: 29, [18F]GE-179; mGluR5: 123, [11C]ABP688), norepinephrine (NET: 77, [11C]MRB), histamine (H3: 8, [11C]GSK189254), cannabinoids (CB1: 77, [11C]OMAR), and opioids (MOR: 204, [11C]Carfentanil). We derived the principal patterns of spatial variance (i.e., gradients) across these 19 maps using principal component analysis (PCA, with each map first scaled and centered), using the *prcomp* and *PCAtest*^*74*^ functions in *R*^*75*^. Permutation testing was used to determine statistically-significant principal components^74^, with *p*-values calculated by comparing the empirical eigenvalue of each component to a null distribution of eigenvalues derived from 1,000 random permutations of the underlying data. We retained the principal components with *p* < .05 for further analysis, as representative topographies of the principal normative neurochemical gradients of the human brain. To test the importance of synapse density, we also retrieved from *neuromaps* the cortical atlas topography of synaptic vesicle glycoprotein 2A (76 adults, [11C]UCB-J).

### Modeling of Spatial Colocalization

We z-scored the individual patient topographies of frequency-specific neurophysiological signal power and cortical thickness with respect to the means and standard deviations of the respective maps of the control group, resulting in cortical maps of alterations from healthy variants for each patient with PD.

We then estimated the alignment of those neurophysiological and structural alterations topographies with the *neuromaps* principal gradients using linear mixed-effects modeling via the *nlme* package in *R*. For each patient, this approach models a nested intercept and slope representing the linear relationship between region-wise cortical alterations and the selected *neuromaps* gradient/atlas, and thus exploits the within-subject variability in the data that would be ignored by group-level colocalization analysis. These models were computed with the following form: *neuromap pc ∼* alterations*(s) + covariates, random = (∼1 +* alterations*(s)* | *participant)*. Initial MEG-*neuromaps* models included all four canonical frequency-band alterations in a single model per each *neuromap gradient*, and alterations exhibiting significant relationships with the same sign were then averaged across frequencies (per each cortical region) for visualization and secondary analysis. For each significant relationship between *neuromap* principal gradients and cortical alteration metrics, we performed post-hoc testing to assess the specificity of the effect to each neurotransmitter system. This was done by regressing (i.e., using a comparable linear mixed-effects model) the relevant averaged multi-frequency alterations on each *neuromap* atlas that exhibited significant loadings onto the relevant neurochemical gradient, with false discovery rate correction across atlases using the Benjamini-Hochberg method.

We mapped the cortical regions contributing most to significant relationships between structural/neurophysiological alterations and *neuromaps* by first extracting the mean (i.e., across individuals) of the mixed-effect model residuals for each region and standardizing these means to the variability (i.e., standard deviation) of the model’s residuals from the same region. Next, the absolute value of these standardized region-wise residuals was computed and subtracted from one. This resulted in one value per region signifying the absolute standardized fit of that region to the overall topographical alignment model, with higher values indicating better fit.

We examined whether significant alignments between the structural/neurophysiological alterations observed in patients and the *neuromaps* principal components are moderated by cognition (i.e., attention, visuospatial function, memory, executive function, and language), self-reported psychiatric symptoms (i.e., depression and/or anxiety), and motor function (i.e., UPDRS-III scores). The effects of continuous moderators (e.g., cognitive and motor function scores) were visualized by (1) extracting and plotting values corresponding to the lower and upper quartile of that moderator and (2) extracting the nested model slope coefficients per participant and plotting these against the moderator. To visualize the cortical regions contributing to significant moderations, we computed an absolute standardized fit metric, similar to the one described above, for the upper and lower quartile of the moderator (or, in the case of psychiatric symptoms, separately for those with and those without reported symptoms) then subtracted these subgroup fits per region.

In addition to age, which was included as a covariate in all instances, we accounted for potential modeling confound effects of disease duration, head motion, eye movements, and heart rate variability by including these factors as nuisance covariates in post-hoc modeling. We also examined whether any of our findings could be accounted for by shifts in the arrhythmic component of the neurophysiological spectrum by testing for colocalization effects between each neurochemical gradient and alterations in the aperiodic slope parameter of *specparam*^69^.

We used the Benjamini-Hochberg method to correct for multiple comparisons across related hypotheses, with a threshold for significance set to *p*_*FDR*_ < .05 across all relevant tests.

We conducted spatial comparisons between group-level maps using general linear models and nonparametric spin-tests with autocorrelation-preserving null models (1,000 Hungarian spins; threshold: *p* < .05)^76^.

## Data Availability

Data used in the preparation of this work are available through the Clinical Biospecimen Imaging and Genetic (C-BIG) repository (https://www.mcgill.ca/neuro/open-science/c-big-repository), the PREVENT-AD open resource (https://openpreventad.loris.ca/), and the OMEGA repository (https://www.mcgill.ca/bic/resources/omega). Normative neurotransmitter density data are available from neuromaps (https://github.com/netneurolab/neuromaps).

https://openpreventad.loris.ca/

https://www.mcgill.ca/bic/resources/omega

https://github.com/netneurolab/neuromaps

## Data Availability

Data used in the preparation of this work are available through the Clinical Biospecimen Imaging and Genetic (C-BIG) repository (https://www.mcgill.ca/neuro/open-science/c-big-repository)^57^, the PREVENT-AD open resource (https://openpreventad.loris.ca/)^59^, and the OMEGA repository (https://www.mcgill.ca/bic/resources/omega)^60^. Normative neurotransmitter density data are available from *neuromaps* (https://github.com/netneurolab/neuromaps)^72^.

## Acknowledgments

This work was supported by grant F32-NS119375 to AIW from the United States National Institutes of Health (NIH); to JDSC as a doctoral fellowship from Natural Science and Engineering Research Council of Canada (NSERC); to EAF as a Foundation Grant from the Canadian Institutes of Health Research (CIHR; FDN-154301) and the CIHR Canada Research Chair (Tier 1) of Parkinson’s Disease; and to SB from by a NSERC Discovery grant, the Healthy Brains for Healthy Lives initiative of McGill University under the Canada First Research Excellence Fund, the CIHR Canada Research Chair (Tier 1) of Neural Dynamics of Brain Systems and the NIH (1R01EB026299). Data collection and sharing for this project was provided by the Quebec Parkinson Network (QPN), the Pre-symptomatic Evaluation of Novel or Experimental Treatments for Alzheimer’s Disease (PREVENT-AD; release 6.0) program, and the Open MEG Archives (OMEGA). The funders had no role in study design, data collection and analysis, decision to publish, or preparation of the manuscript.

The QPN is funded by a grant from Fonds de recherche du Québec - Santé (FRQS). PREVENT-AD was launched in 2011 as a $13.5 million, 7-year public-private partnership using funds provided by McGill University, the FRQS, an unrestricted research grant from Pfizer Canada, the Levesque Foundation, the Douglas Hospital Research Centre and Foundation, the Government of Canada, and the Canada Fund for Innovation. Private sector contributions are facilitated by the Development Office of the McGill University Faculty of Medicine and by the Douglas Hospital Research Centre Foundation (http://www.douglas.qc.ca/). OMEGA and the Brainstorm app are supported by funding to SB from the NIH (R01-EB026299), a Discovery grant from the Natural Science and Engineering Research Council of Canada (436355-13), the CIHR Canada research Chair in Neural Dynamics of Brain Systems, the Brain Canada Foundation with support from Health Canada, and the Innovative Ideas program from the Canada First Research Excellence Fund, awarded to McGill University for the HBHL initiative.

