## Supplemental Information for "Structural and neurophysiological alterations in Parkinson’s disease are aligned with cortical neurochemical systems"

Alex I. Wiesman* et al.

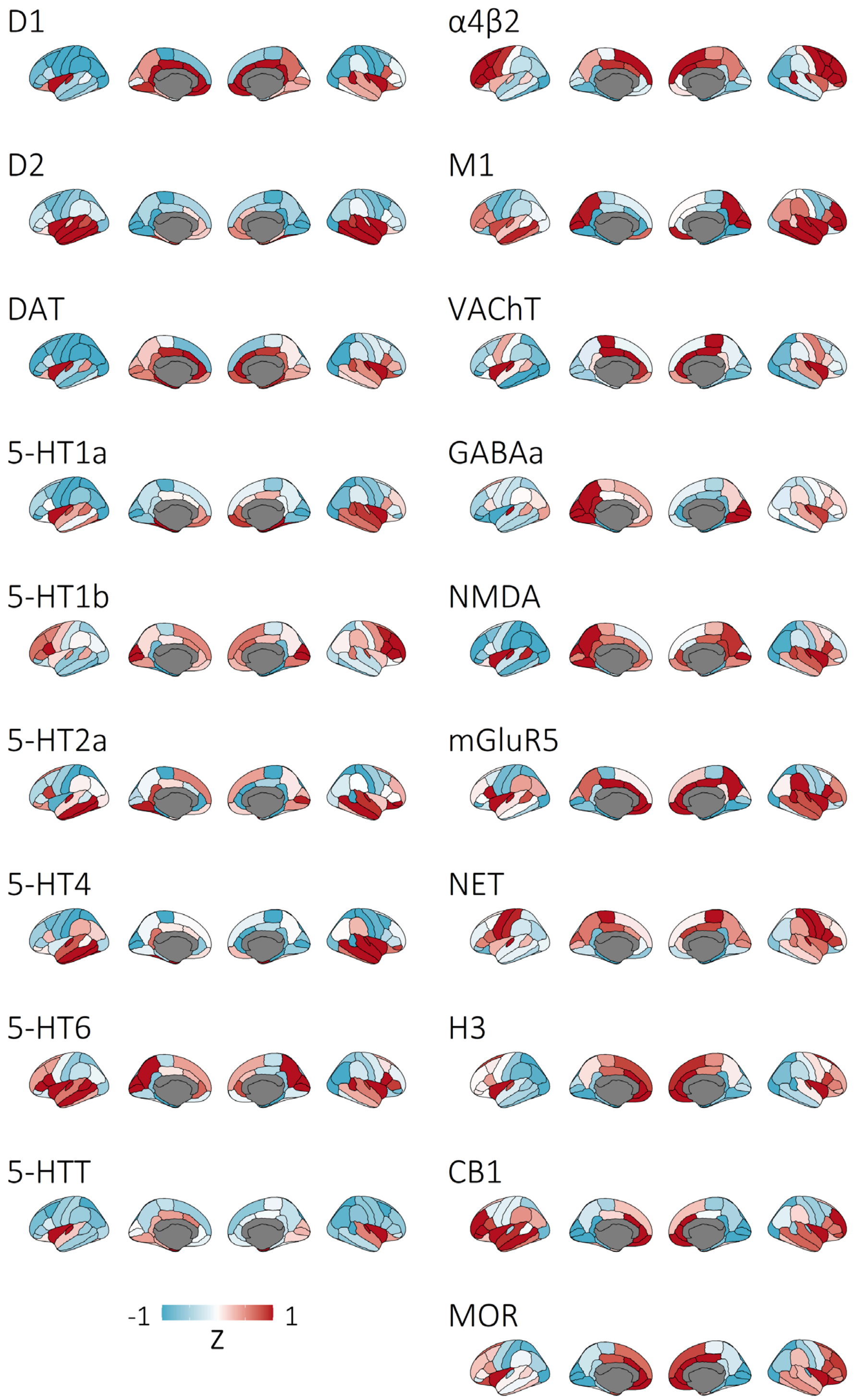


**Figure S1. Normative atlases of 19 neurotransmitter system densities.** Brain maps represent normative densities of 19 neurotransmitter receptor and transporters from the *neuromaps* multi-atlas, measured with positron emission tomography (PET) in healthy adult participants. Maps have been internally z-scored for comparison of spatial patterns across PET outcome measures with different scales.

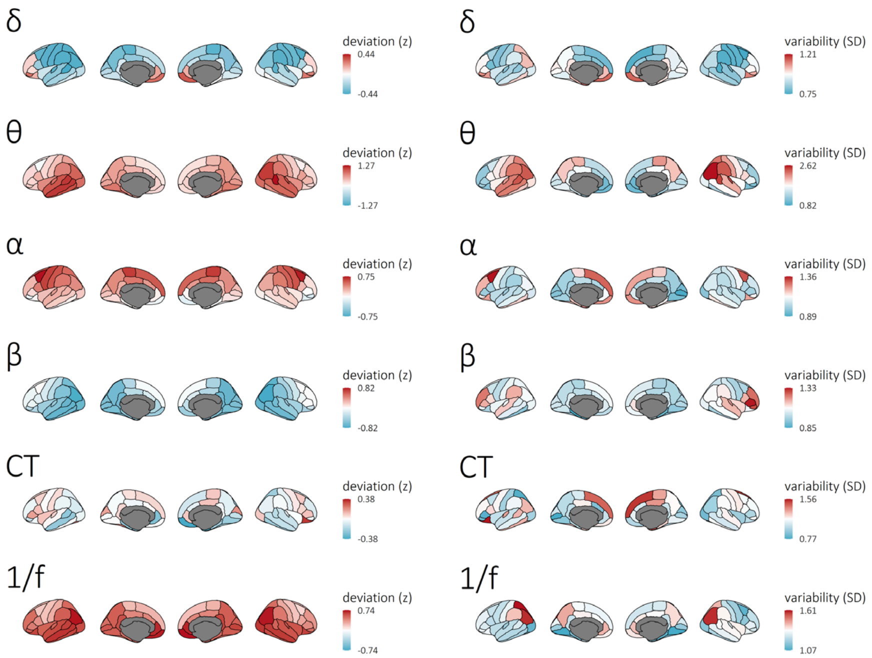


**Figure S2. Central tendency and variability of cortical neurophysiological alterations in patients with Parkinson's disease.** Brain maps in the left column display the mean deviation of each indicated metric relative to healthy controls, derived across all patients with Parkinson's disease. Maps in the right column indicate the variability of these deviations across patients, which was leveraged by the linear mixed model approach to colocalization analysis.

**Table S1.** Demographic comparisons and patient group clinical profile – neurophysiological data.

| Group | **Age** (years) | **Sex** (% male) | **Handedness** (# left/ambi) | **Education** (years) |
|---|---|---|---|---|
| **HC** (N = 65) | 63.02 (8.13) | 64.62 | 3/3 | 15.85 (3.72)* |
| **PD** (N = 79) | 64.60 (8.21) | 70.89 | 5/3 | 15.05 (3.04) |
| *p* | .382 | .422 | .883 | .444 |
| | **MoCA** (N = 70) | **UPDRS-III** (N = 61) | **Hoehn & Yahr** (N = 57) | **% Taking DA Agonists** (N = 66) |
| **PD** | | | | |
| **Range** | 12 – 30 | 7 – 71 | 1 – 3 | – |
| **Mean** (SD) | 24.39 (3.98) | 32.44 (14.64) | 1.90 (0.70) | 31.82 |

HC: healthy control group; PD: Parkinson's disease group; MoCA: Montreal Cognitive Assessment; UPDRS-III: Unified Parkinson's Disease Rating Scale part III; DA: Dopamine. Unless otherwise indicated, values indicate means and associated parentheticals indicate standard deviations. P-values indicate significance of between-groups Mann-Whitney U tests and chi-square tests for continuous and categorical variables, respectively. *N = 59; highest level of education was not reported by six HC participants.

**Table S2.** Demographic comparisons and patient group clinical profile – cortical thickness data.

| Group | Age (years) | Sex (% male) | Handedness (# left/ambi) | Education (years) |
|---|---|---|---|---|
| **HC** (N = 37) | 64.76 (9.54) | 48.65 | 2/0* | 14.71 (4.66)** |
| **PD** (N = 65) | 64.68 (7.93) | 67.69 | 3/2 | 14.76 (2.57) |
| *p* | .656 | .058 | .593 | .395 |
| | **MoCA** (N = 57) | **UPDRS-III** (N = 51) | **Hoehn & Yahr** (N = 50) | **% Taking DA Agonists** (N = 55) |
| **PD** | | | | |
| **Range** | 12 – 30 | 7 – 71 | 1 – 3 | – |
| **Mean** (SD) | 24.16 (4.04) | 32.63 (14.19) | 1.89 (0.72) | 30.91 |

HC: healthy control group; PD: Parkinson's disease group; MoCA: Montreal Cognitive Assessment; UPDRS-III: Unified Parkinson's Disease Rating Scale part III; DA: Dopamine. Unless otherwise indicated, values indicate means and associated parentheticals indicate standard deviations. P-values indicate significance of between-groups Mann-Whitney U tests and chi-square tests for continuous and categorical variables, respectively. *N = 17; handedness was not reported by twenty HC participants. **N = 28; highest level of education was not reported by nine HC participants.

### *Consortium Authors*

A complete listing of **PREVENT-AD Research Group** can be found in the PREVENT-AD database:

https://preventad.loris.ca/acknowledgements/acknowledgements.php?date=[2022-02-01]

The members of the **Quebec Parkinson Network** are listed alphabetically below:


Isabelle Beaulieu-Boire - Université de Sherbrooke, Sherbrooke, Canada
Pierre Blanchet - Université de Montréal, Montréal, Canada
Sarah Bogard - Montreal Neurological Institute, McGill University, Montreal, Canada
Manon Bouchard – Clinique NeuroLévis, Lévis, Canada
Sylvain Chouinard - Université de Montréal, Montréal, Canada
Francesca Cicchetti - CHU de Québec-Université Laval, Québec, Canada
Martin Cloutier - Neuro-Rive-Sud, Longueuil, Canada
Alain Dagher - Montreal Neurological Institute, McGill University, Montreal, Canada
Samir Das - Montreal Neurological Institute, McGill University, Montreal, Canada
Clotilde Degroot - Montreal Neurological Institute, McGill University, Montreal, Canada
Alex Desautels - Université de Montréal, Montréal, Canada
Marie Hélène Dion - Université de Montréal, Montréal, Canada
Janelle Drouin-Ouellet - Université de Montréal, Montréal, Canada
Anne-Marie Dufresne - CHU de Québec-Hôpital de l'Enfant-Jésus, Québec, Canada
Nicolas Dupré - CHU de Québec-Université Laval, Québec, Canada
Antoine Duquette - Université de Montréal, Montréal, Canada
Thomas Durcan - Montreal Neurological Institute, McGill University, Montreal, Canada
Lesley K. Fellows - Montreal Neurological Institute, McGill University, Montreal, Canada
Edward Fon - Montreal Neurological Institute, McGill University, Montreal, Canada
Jean-François Gagnon - Université du Québec à Montréal, Montréal, Canada
Ziv Gan-Or - Montreal Neurological Institute, McGill University, Montreal, Canada
Angela Genge - Montreal Neurological Institute, McGill University, Montreal, Canada
Nicolas Jodoin - Centre hospitalier de l'Université de Montréal, Montréal, Canada
Jason Karamchandani - Montreal Neurological Institute, McGill University, Montreal, Canada
Anne-Louise Lafontaine - Montreal Neurological Institute, McGill University, Montreal, Canada
Mélanie Langlois - CHU de Québec-Université Laval, Québec, Canada
Etienne Leveille - Montreal Neurological Institute, McGill University, Montreal, Canada
Martin Lévesque - Université Laval, Québec, Canada
Calvin Melmed - McGill University, Montreal, Canada
Oury Monchi - Université de Montréal, Montréal, Canada
Jacques Montplaisir - Université de Montréal, Montréal, Canada
Michel Panisset - Université de Montréal, Montréal, Canada
Martin Parent - Université Laval, Québec, Canada
Minh-Thy Pham-An - Hôpital Hôtel-Dieu de Saint-Jérôme, Saint-Jérôme, Canada
Jean-Baptiste Poline - Montreal Neurological Institute, McGill University, Montreal, Canada
Ronald Postuma - Montreal Neurological Institute, McGill University, Montreal, Canada

Emmanuelle Pourcher - CHU de Québec-Université Laval, Québec, Canada
Trisha Rao - McGill University, Montreal, Canada
Jean Rivest - CHU de Québec-Université Laval, Québec, Canada
Guy Rouleau - Montreal Neurological Institute, McGill University, Montreal, Canada
Madeleine Sharp - Montreal Neurological Institute, McGill University, Montreal, Canada
Valérie Soland - CHU de Montréal, Montréal, Canada
Michael Sidel - Montreal Neurological Institute, McGill University, Montreal, Canada
Sonia Lai Wing Sun - Montreal Neurological Institute, McGill University, Montreal, Canada
Alexander Thiel - McGill University, Montreal, Canada
Paolo Vitali - McGill University, Montreal, Canada
